# Place of Death in Ventricular Arrhythmias in the United States: A 25-Year Population-Based Analysis From 1999 2024

**DOI:** 10.64898/2026.01.14.26344154

**Authors:** Muhammad Abdullah Naveed, Muhammad Junaid Razzak, Muhammad Hasan, Ahila Ali, Muhammad Omer Rehan, Siddharth Agarwal, Sivaram Neppala, Abhishek Deshmukh, Christopher V DeSimone, Muhammad Bilal Munir

## Abstract

**Background:** Ventricular arrhythmias (VAs) are a proximate mechanism of sudden cardiac death, yet national patterns in place of death (POD) and their determinants remain sparsely described. We quantified 25-year trends and factors associated with POD among UAs decedents in the United States.

**Methods:** We analyzed CDC WONDER Multiple Cause of Death data (1999–2024) for adults ≥25 years with ventricular arrhythmias (ICD-10 I47.2, I49.0) as underlying cause. POD was categorized as inpatient, outpatient/emergency department (ED), home, hospice/nursing, or other/unknown. Covariates included age, sex, race, Hispanic origin, and urbanization. We calculated Annual and Average Annual Percent Changes (AAPCs and APC) using Age-Adjusted Mortality Rates (AAMRs), and fit multinomial logistic regression (reference = inpatient) to obtain adjusted odds ratios (ORs, 95% CIs).

**Results:** Among 433,988 ventricular arrhythmia (VA) deaths, POD was inpatient 62.0%, outpatient/ED 18.1%, home 11.1%, hospice/nursing 5.8%, other 3.1%. Inpatient deaths increased from 57.8% (1999) to 66.1% (2024). AAMRs declined sharply from 13.3 per 100,000 in 1999 to 6.3–6.5 during 2010–2019, then rose to 7.4 in 2021 and fell to 6.8 in 2024. In home vs inpatient: ≥85 years, medium/small metropolitan counties and rural counties had higher odds of VA deaths, whereas younger age groups, females, Black, American Indian, Asian/Pacific Islander individuals and Hispanic individuals had lower odds. In outpatient/ED vs inpatient: 25–44 years, 45–64 years, males, Black, American Indian and Asian/Pacific Islander individuals had higher odds, whereas ≥85 years and females had lower odds. In hospice/nursing facilities: ≥85 years, females, Whites, non-Hispanic individuals, medium/small metropolitan counties and rural counties had higher odds of VA deaths, whereas younger age groups, Black, American Indian, Asian/Pacific Islander individuals and Hispanic individuals had markedly lower odds.

**Conclusion:** From 1999–2024, VA deaths shifted toward hospitals. Persistent disparities by age, sex, race/ethnicity, and rurality highlight the need to expand equitable advance care planning and device deactivation discussions.

## Introduction

Ventricular arrhythmias (VAs) principally sustained ventricular tachycardia and ventricular fibrillation, are a proximate mechanism of sudden cardiac death (SCD) and an important determinant of cardiovascular mortality [1,2]. Epidemiologically, the burden is heterogenous, incidence rises steeply with age, remains consistently higher in men than in women, and disproportionately affects Black individuals [3], Globally, SCD accounts for an estimated 4 to 5 million deaths annually [4], and approximately 350,000 deaths occur due to SCD every year, with VAs as the predominant participants [5,6].

VAs arise from the interaction of arrhythmogenic substrate and precipitating triggers in vulnerable ventricular myocardium. Structural remodeling, particularly post-infarct scar and border-zone fibrosis, produces conduction anisotropy and dispersion of refractoriness, creating conduction channels that sustain re-entrant wavefronts; in parallel, ischemia, inflammatory signaling, and disordered intracellular calcium flux promote triggered firing and enhanced automaticity [7–9]. Once established, sustained tachyarrhythmia compromises coordinated ventricular contraction, rapidly reduces stroke volume and perfusion, and often degenerates into fibrillation, resulting in cessation of cardiac output. In the absence of immediate defibrillation, this cascade advances within minutes to global myocardial ischemia, circulatory collapse, and death [7–9].

VAs impose a substantial financial burden on healthcare systems, driven largely by emergency response demands, intensive hospitalization, device implantation, and long-term management of post-cardiac arrest patients. Across the United States, the financial expenditures per annum attributable to cardiac arrest approximates at $33 billion, emphasizing the considerable resource utilization inherent to resuscitation and post-arrest care [10].

VAs-related mortality varies substantially across settings due to the probability of immediate recognition, defibrillation, and advanced life support varying across home, public, and hospital settings. Delayed bystander response, restricted access to automated external defibrillators, and fragmentation in post-resuscitation care intensify these disparities. Despite these evident setting-specific determinants, longitudinal analysis delineating VAs-specific mortality trends by death settings remain sparse, damping data-driven targeting of interventions such as community defibrillation timing and in-hospital rescue optimization.

To address this deficit, the current investigation draws on national mortality data of US adults aged ≥25 years from the CDC WONDER Multiple Cause of Death database, analyzing 1999–2024 trends in VAs-related deaths across distinct care environments. Deaths were stratified into five operational settings, (1) inpatient facilities, (2) outpatient or emergency departments, (3) homes, (4) hospice or long-term care institutions, and (5) other/unknown locations categorized by CDC WONDER database. By integrating temporal and contextual dimensions, this approach links mechanistic insight with real-world outcomes, enabling clinicians and health-system planners to channel resources and quality-improvement efforts toward the settings where timely intervention can yield the maximum survival gains.

## Methods

### Data Source and Study Population

We conducted a retrospective analysis of national mortality data using the CDC WONDER (Wide-ranging Online Data for Epidemiologic Research) Underlying Cause of Death database for the years 1999-2024 (11). This publicly available resource contains deidentified death certificate information for U.S. residents, including underlying cause coded by ICD-10. We identified all decedents aged ≥25 years with an underlying cause of death or contributing cause of death of VAs, defined by ICD-10 code I47.2 and I49.0 (12). We excluded any deaths due to external or non-natural causes, as indicated by ICD-10 codes V01-X59, X60-X84, X85-Y09 (injury, poisoning, and external causes) or R00-R99 (symptoms, signs, and ill-defined conditions). Deaths with missing or unknown place of death were excluded from regression analyses. In accordance with CDC guidance, counts or rates based on small numbers (fewer than 10 deaths) were suppressed to protect confidentiality.

This study was deemed exempt from institutional review board (IRB) review because it used publicly available, deidentified data and did not involve human subjects research. Study reporting followed STROBE (Strengthening the Reporting of Observational Studies in Epidemiology) guidelines (13).

### Measures

Outcome (Place of Death): The primary outcome was place of death, as reported on the death certificate. CDC WONDER categorizes place of death into several mutually exclusive fields (e.g., inpatient medical facility, outpatient or emergency department, decedent’s home, hospice facility, nursing home/long-term care, other, unknown) (14). For this analysis we collapsed these into five categories: (1) Inpatient medical facility (hospital), (2) Outpatient facility or emergency department, (3) Home, (4) Hospice facility or nursing home/long-term care, and (5) Other/unknown. The “Other/unknown” category comprised deaths reported as “dead on arrival,” those that occurred in other specified places (e.g., commercial buildings, public grounds, or in transit), and those for which the place of death was not stated or was unknown. We combined these due to the small number of deaths in each sub-category when analyzed separately. Category assignments followed CDC conventions (14).

Independent Variables: We analyzed demographic and geographic covariates available on the death certificate. Age at death was categorized as 25-44, 45-64, 65-84, and ≥85 years. Sex (male, female) and races (Hispanic, White, Black, Other race) were reported as recorded on the certificate. Hispanic ethnicity was coded as yes or no (Hispanic origin). Urbanization of the decedent’s residence was classified using the 2013 National Center for Health Statistics (NCHS) Urban-Rural Classification Scheme for Counties (15). In brief, counties are assigned to all the available six levels of urbanization in the CDC WONDER database, which have been grouped into four categories: large metropolitan, medium metropolitan, small metropolitan, and nonmetropolitan rural (15).

### Statistical Analysis

We first summarized the distribution of place of death by calendar year and by demographic subgroup. Descriptive statistics included counts and proportions of decedents in each place-of-death category, stratified by year, age group, sex, race/ethnicity, and urbanization level. We examined temporal trends in the proportion of deaths occurring in each setting using graphical methods (plots of annual proportions) and assessed linear trend by treating calendar year as a continuous variable.

To estimate associations between decedent characteristics and place of death, we fit a multinomial logistic regression model with place of death as the outcome (16). In this model the reference category was inpatient medical facility. We included all covariates simultaneously (age group, sex, race, Hispanic ethnicity, and urbanization) to estimate adjusted odds ratios (ORs) and 95% confidence intervals (CIs) for dying in each nonreference setting (home, hospice/nursing home, outpatient/ER, and other) relative to dying in an inpatient facility. Because the model included all predictors at once, the ORs represent the independent association of each variable with place of death. All statistical tests were two-sided with a significance level of α = 0.05; P values <.05 were considered statistically significant.

All analyses were performed in RStudio (version 4.3.2; R Foundation for Statistical Computing, Vienna, Austria) [17]. Summary tables report adjusted proportions and differences in proportions with 95% CIs. Regression results are presented as adjusted ORs with 95% CIs. We used standard routines for multinomial logistic regression and trend tests in R using appropriate statistical packages.

## Results

### Overview of total VA deaths and place of death

From 1999 through 2024, a total of 433,988 U.S. adults (≥25 years) died with ventricular arrhythmias (VA) as the underlying cause **(Table 1)**. Most of the deaths occurred in inpatient hospital 268,965 (62.0%), followed by outpatient/ED 78,536 (18.1%), home 47,959 (11.1%), hospice or nursing facility 25,206 (5.8%), and other 13,322 (3.1%) **(as shown in Table 1, Figure 2 and 4)**.

**Table 1:**
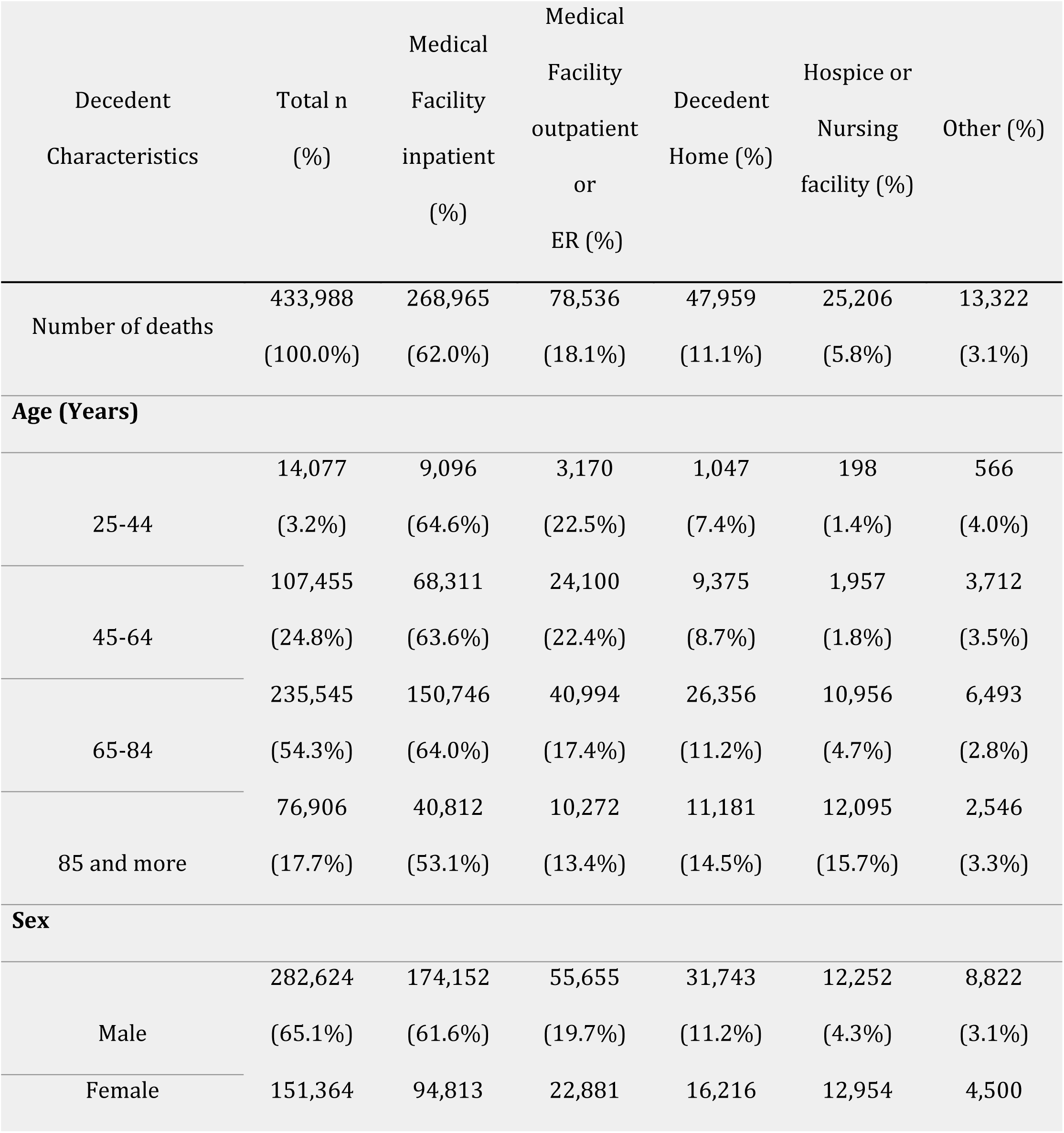
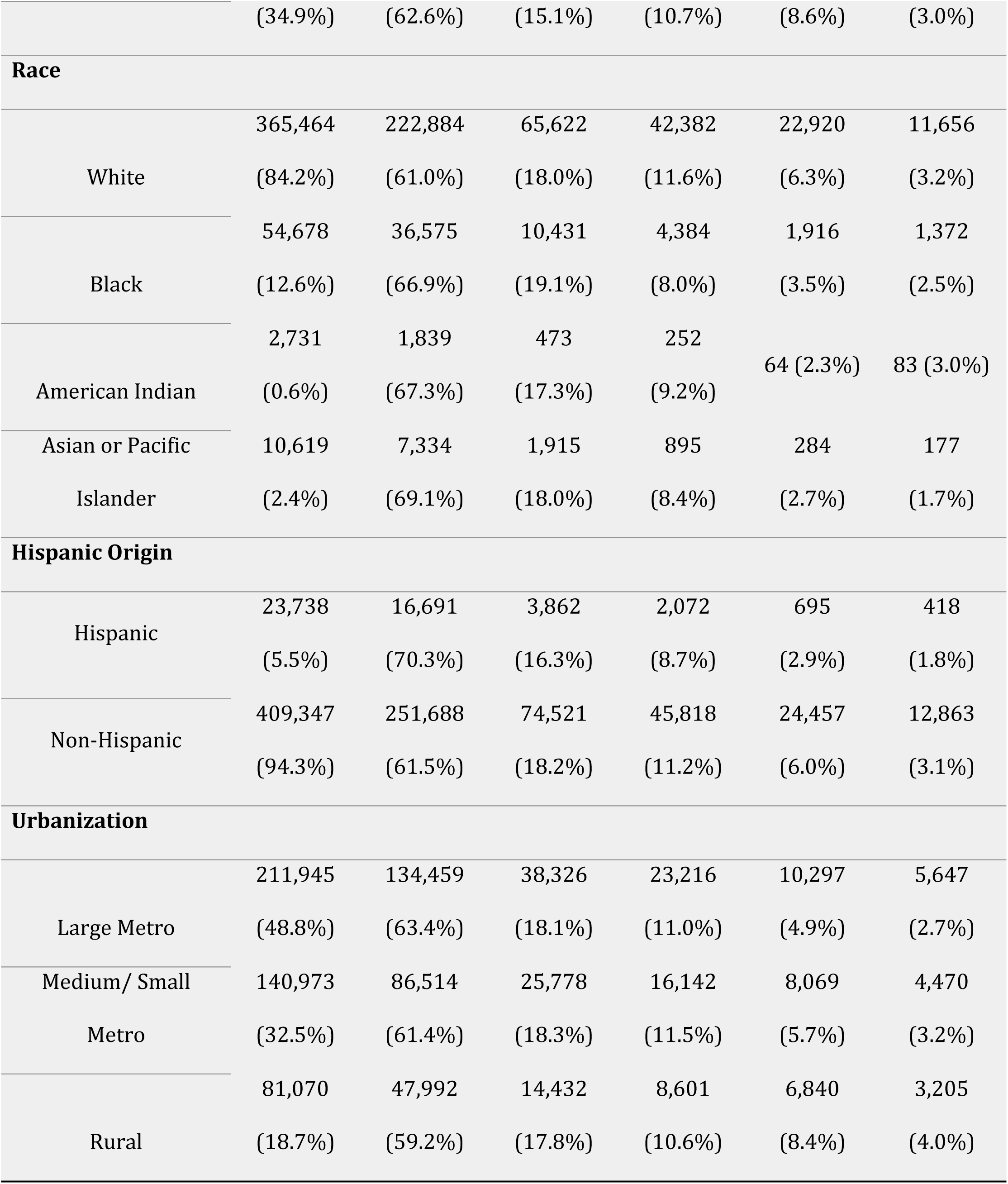
Aggregated Data for Adults for places of death by decedent characteristics for Ventricular Arrhythmias (1999-2024)

### Decedents’ characteristics

Deaths were concentrated in the aging population: 65–84 years accounted for 54.3% of VA deaths, and an additional 17.7% occurred among those ≥85 years; in parallel, 45–64 years comprised 24.8%, and 25–44 years just 3.2% **(Figure 2A)**. Men accounted for 65.1% of deaths, substantially more than women (34.9%) **(Figure 2B)**. Most descendants were White (84.2%), with Black (12.6%), Asian/Pacific Islander (2.4%), and American Indian (0.6%) minorities. Hispanic origin represented 5.5% of VA deaths **(Figure 2C)**. Geographically, nearly half of deaths occurred among residents of large metropolitan counties (48.8%), followed by medium/small metros (32.5%) and rural areas (18.7%) **(as shown in Table 1**, **Figure 2D and 4)**.

### Trends in age-adjusted mortality over time

Age-adjusted VA mortality declined sharply during 1999–2007 [APC −8.51% (95% CI, −8.98 to −8.14); p < 0.001], followed by a non-significant near-plateau till 2018 [APC −0.28% (95% CI, −0.94 to 0.06); p = 0.08]. Rates then increased from 2018 to 2021 [APC +5.49% (95% CI, +2.72 to +6.83); p = 0.006] and declined again till 2024 [APC −2.65% (95% CI, −5.59 to −1.04); p = 0.0132]. Anchor values from the annual tables: 13.3 per 100,000 in 1999, 6.3–6.5 during 2010–2019, 6.9 in 2020, 7.4 in 2021, and 6.8 in 2024 **(as shown in Figure 1A and Supplemental Table 1).**

**Figure 1A & 1B:**
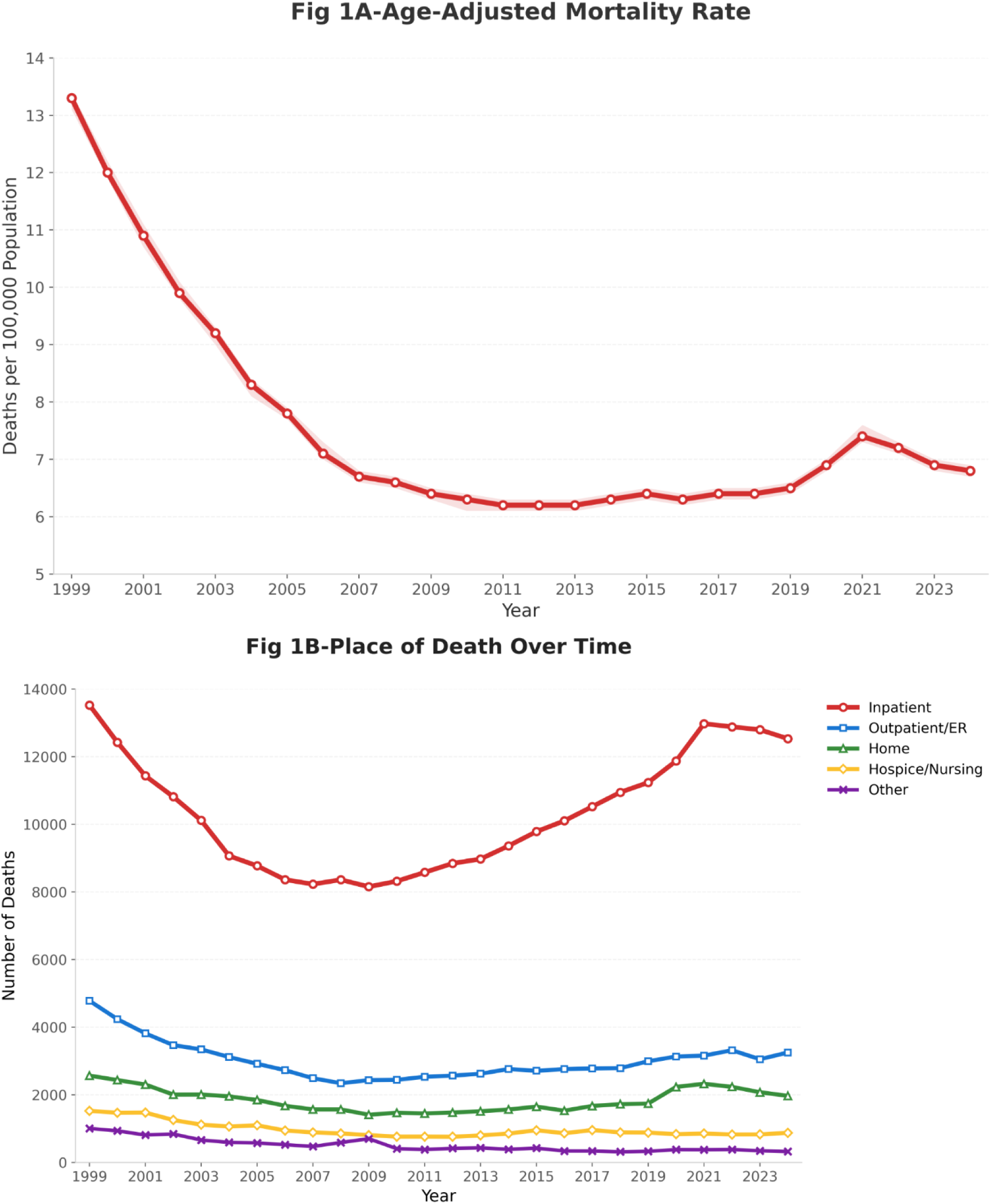
Trend and Number of Deaths According to Places of Deaths related to Ventricular Arrhythmia from 1999 - 2024.

### Changes in place-of-death distribution over time

Tracking the joinpoint years, the share of deaths occurring in hospital rose from 57.8% in 1999 to 60.3% in 2007, then to 65.7% by 2018; it remained high during 2021 (65.9%) and 2024 (66.1%). Outpatient/ED deaths decreased from 20.4% (1999) to 18.2% (2007) and 16.7% (2018), dipped to 16.0% (2021), and edged to 17.2% (2024). Home deaths remained near 11% in the early segments, 11.0% (1999) and 11.5% (2007), then dipped to 10.3% (2018), rose during the pandemic to 11.8% (2021), and dropped to 10.4% in 2024. Hospice/nursing facility deaths declined throughout the period, 6.5% in 1999 and 2007, 5.3% (2018), 4.3% (2021), and 4.6% in 2024. Deaths in other locations also contracted from 4.3% in 1999 to 3.5% in 2007, 1.9% (2018–2021), and 1.7% in 2024. **(as shown in Figure 1B and Supplemental Table 2)**.

### Demographic variation in place of death

#### Age

Among 25–44 years, deaths were concentrated in medical facility/inpatient (64.6%) with very few in hospice/nursing (1.4%). In 45–64 years, medical facility/inpatient accounted for the highest number of deaths (63.6%), and least deaths occurred in hospice/nursing (1.8%). In 65–84 years, the highest death toll was observed in medical facility/inpatient (64%), with the least number of deaths observed in other settings (2.8%). Among those ≥85 years, medical facility/inpatient and other places accounted for most and least fatalities (53.1% and 3.3%, respectively) **(as shown in Table 1**, **Figure 2A and 4)**.

**Figure 2A, 2B, 2C & 2D:**
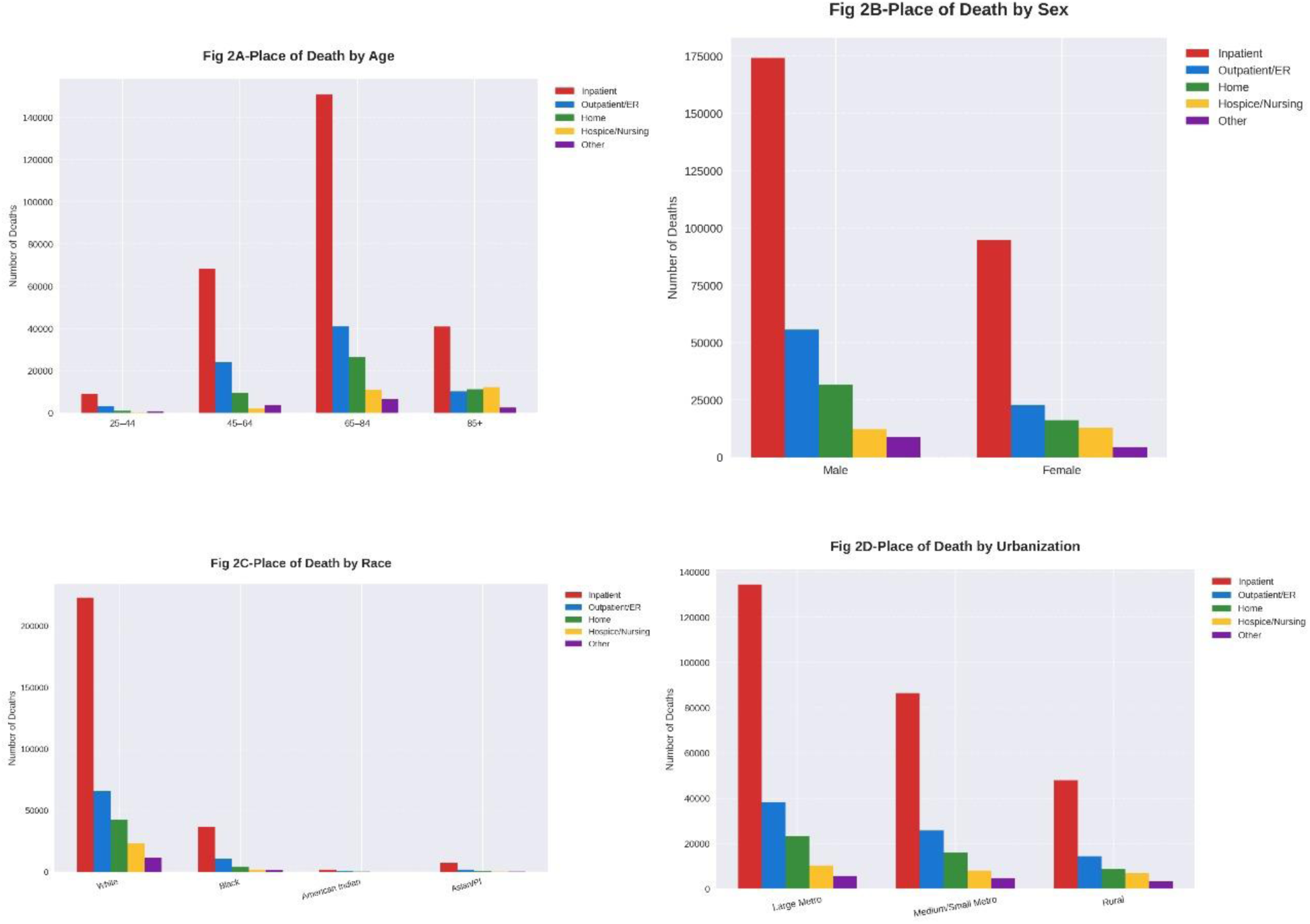
Percentage of Deaths Stratified by Age Group, Sex, Race, and Urbanization Level According to Place of Death Among Ventricular Arrhythmia Decedents, 1999–2024.

#### Sex

Across sex strata, medical facility/inpatient remained the predominant setting (male: 61.6%; Female: 62.6%), whereas other settings accounted for least deaths (male: 3.1%; Female: 3%) **(as shown in Table 1**, **Figure 2B and 4)**.

#### Race

Racial gradients were modest but consistent. Medical facility/inpatient was the dominant setting across all groups (Asian/Pacific Islanders: 69.1%; American Indians: 67.3%; Black individuals: 66.9%; Whites: 61.0%). In contrast, other places accounted for the least number of deaths in Whites (3.2%), Black individuals (2.5%), and Asian or Pacific Islanders (1.7%). In contrast, hospice or nursing was the uncommon location of death for American Indians (2.3%) **(as shown in Table 1**, **Figure 2C and 4)**.

#### Hispanic origin

Medical facility/inpatient dominated among Hispanic (70.3%) compared with non-Hispanic (61.5%) descendants. Deaths in other places were less common in both origins (Hispanic: 1.8%; Non-Hispanic: 3.1%) **(as shown in Table 1**, **Figure 4)**.

#### Urbanization

Medical facility/inpatient was the dominant setting in large metropolitan areas (63.4%) and medium/small metros (61.4%), tapering in rural counties (59.2%). Whereas other location deaths were less common (large metropolitan areas: 2.7%; medium/small metros: 3.2%; rural counties: 4%) **(as shown in Table 1**, **Figure 2D and 4)**.

All comparisons across demographic characteristics were statistically significant (p < 0.001 for all), as shown in **Supplemental Table 3**

### Odds ratios for place of death

Odds are expressed as comparison vs referent; reference categories were age 65–84 years, male, White, Non-Hispanic, and large-metropolitan counties.

#### Home vs inpatient

relative to 65–84 years, ≥85 had higher odds (OR = 1.57; 95% CI, 1.53–1.61; p < 0.001), whereas 25–44 and 45–64 had lower odds (OR = 0.66; 95% CI, 0.62–0.70 and OR = 0.78; 95% CI, 0.77–0.81; both p < 0.001). Females had slightly lower odds than males (OR = 0.94; 95% CI, 0.92–0.96; p < 0.001). Compared with Whites, Black (OR = 0.63; 95% CI, 0.61–0.65), American Indian (OR = 0.72; 95% CI, 0.63–0.82), and Asian/Pacific Islander (OR = 0.64; 95% CI, 0.60–0.69) decedents had lower odds of home death (all p < 0.001). Hispanic origin was associated with lower odds than Non-Hispanic (OR = 0.68; 95% CI, 0.65–0.71; p < 0.001). By urbanization, medium/small metro residents had slightly higher odds than large-metro (OR = 1.08; 95% CI, 1.06–1.10; p < 0.001), and rural residents had a smaller but still higher odds (OR = 1.04; 95% CI, 1.01–1.07; p = 0.007). **(as shown in Figure 3A and Supplemental Table 5)**

**Figure 3A, 3B & 3C:**
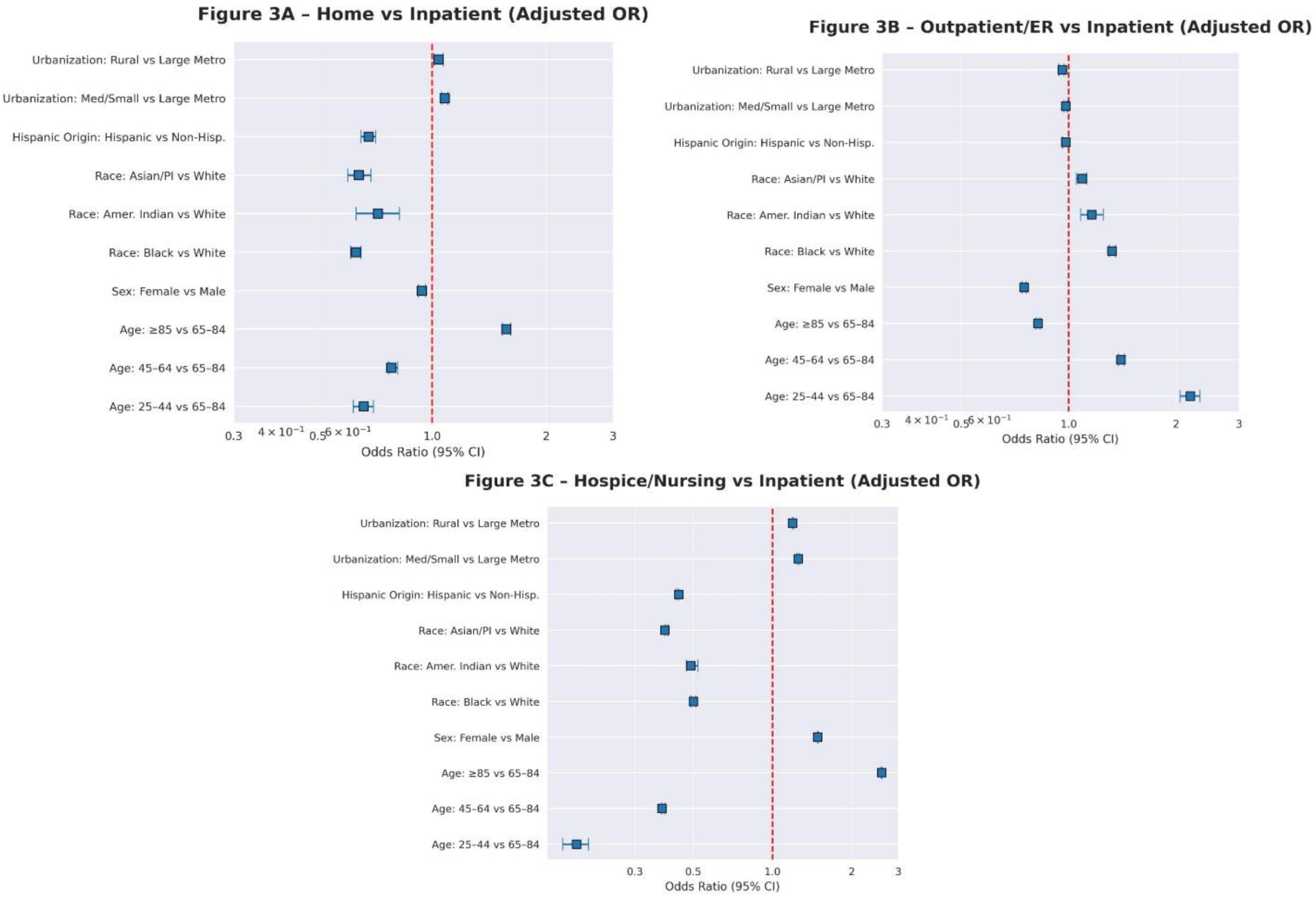
Adjusted Odds Ratios for Place of Death vs Inpatient Deaths in Ventricula Arrhythmia, 1999–2024.

**Figure 4:**
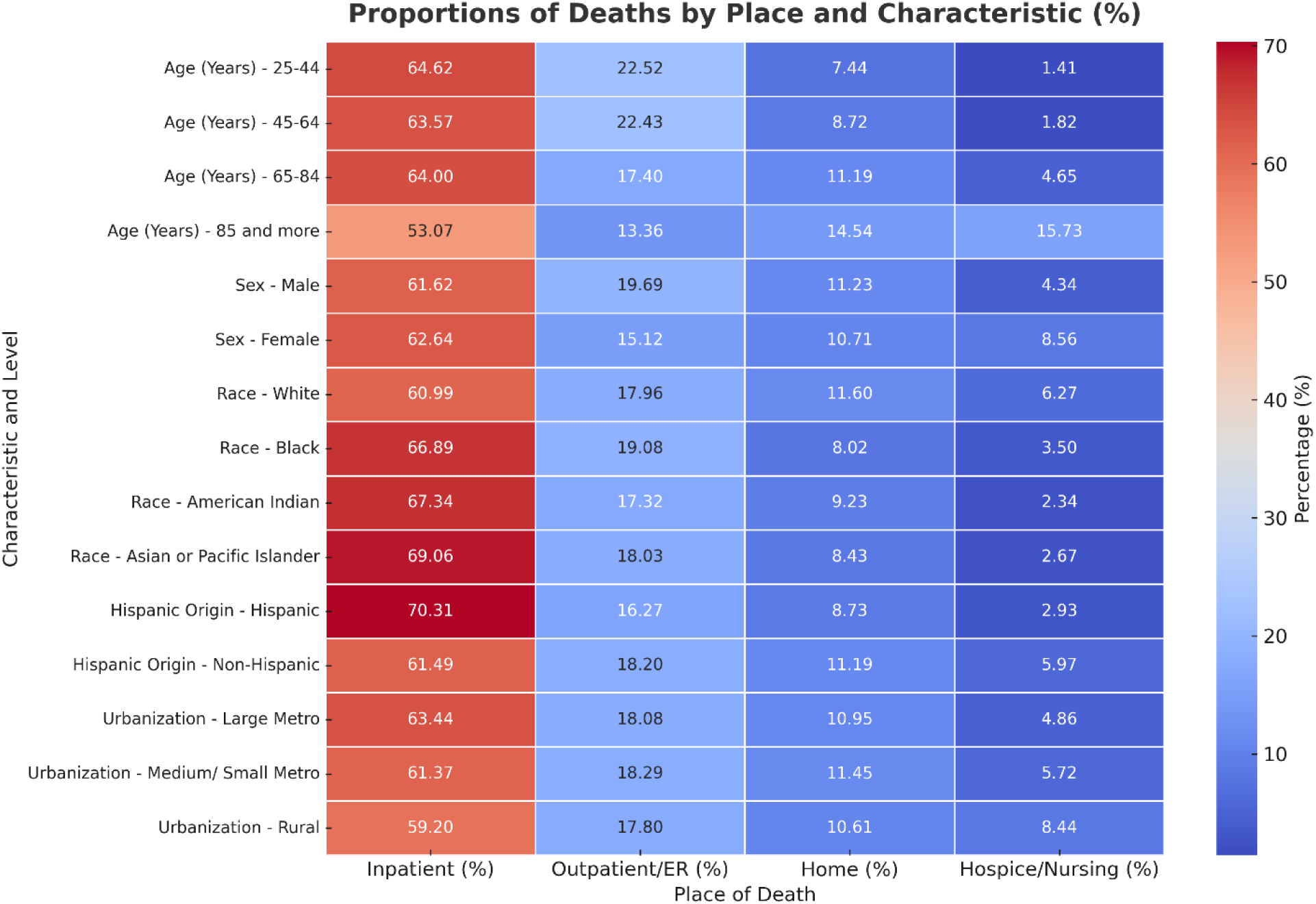
Proportions of Ventricular Arrhythmia Deaths by Place of Death and Demographic Characteristics, 1999–2024.

#### Outpatient/ED vs inpatient

25–44 and 45-64 years were more likely to die in outpatient/ED settings than adults aged 65–84 years (OR = 2.19; 95% CI, 2.05–2.33 and OR = 1.40; 95% CI, 1.38–1.43; both p < 0.001), while ≥85 had lower odds (OR = 0.82; 95% CI, 0.81–0.83; p < 0.001). Females were less likely to die in outpatient/ED settings than males (OR = 0.75; 95% CI, 0.74–0.76; p < 0.001). Relative to Whites, Black (OR = 1.32; 95% CI, 1.30–1.35; p < 0.001), American Indian (OR = 1.16; 95% CI, 1.08–1.25; p < 0.001), and Asian/Pacific Islander (OR = 1.09; 95% CI, 1.05–1.12; p < 0.001) had higher chances of dying in outpatient/ED settings. Hispanic vs non-Hispanic showed a non-significant difference (OR = 0.98; 95% CI, 0.96–1.00; p = 0.081). Compared with large-metro, medium/small metro (OR = 0.98; 95% CI, 0.97–0.99; p = 0.002) and rural (OR = 0.96; 95% CI, 0.94–0.97; p < 0.001) had slightly lower odds. **(as shown in Figure 3B and Supplemental Table 5)**.

#### Hospice/nursing vs inpatient

very old age strongly increased the odds: ≥85 year old had higher odds of dying in hospice/nursing than 65–84 years (OR = 2.59; 95% CI, 2.58–2.60; p < 0.001), whereas 25–44 (OR = 0.18; 95% CI, 0.16–0.20; p < 0.001) and 45–64 (OR = 0.38; 95% CI, 0.38–0.39; p < 0.001) had much lower odds. Females had higher odds than males (OR = 1.48; 95% CI, 1.48–1.49; p < 0.001). Relative to Whites, Black (OR = 0.50; 95% CI, 0.49–0.51; p < 0.001), American Indian (OR = 0.49; 95% CI, 0.47–0.52; p < 0.001), and Asian/Pacific Islander (OR = 0.39; 95% CI, 0.39–0.40; p < 0.001) had lower chances of dying in hospice/nursing homes, and Hispanic origin was associated with lower odds than Non-Hispanic (OR = 0.44; 95% CI, 0.43–0.44; p < 0.001). Versus large-metro, both medium/small metro (OR = 1.25; 95% CI, 1.24–1.26; p < 0.001) and rural (OR = 1.19; 95% CI, 1.19–1.20; p < 0.001) residents had higher odds of hospice/nursing death. **(as shown in Figure 3C and Supplemental Table 5)**.

Standardized mortality ratios stratified by demographic characteristics and place of death are presented in **Supplemental Table 4.**

## Discussion

This 25-year national analysis reveals a defining and countervailing trend in the landscape of mortality from ventricular arrhythmias in the United States: against a backdrop of a substantial decline in overall VA mortality, the hospital has consolidated its role as the dominant site of death, with over 66% of all VA fatalities now occurring in an inpatient setting. This pattern is essentially the reverse of national trends observed for most other chronic illnesses and underscores the singular, precipitous profile of sudden cardiac death (18). Additionally, our findings reveal pronounced and persistent inequities with racial and ethnic minority groups were significantly less likely to die at home or in hospice, highlighting systemic shortcomings in the availability, accessibility, or utilization of advance care planning and palliative care pathways (19,20,21).

The central observation of our analysis is the sustained and intensifying concentration of VA deaths within hospital settings. This trend stands in sharp opposition to the well-established national shift towards home and hospice deaths observed in conditions like heart failure, cerebrovascular diseases, and chronic kidney disease (18,22). Rather than representing a failure of health systems, this divergence is best understood as a direct expression of VA’s core pathophysiology and the clinical frameworks that have been constructed to manage it. The sharp decline in age-adjusted VA mortality from 1999 to 2007, likely attributable to the widespread adoption of evidence-based medical therapies for coronary disease and heart failure and the expanding indications and utilization of implantable cardioverter-defibrillators (ICDs), has been a landmark achievement in cardiovascular medicine (23,24,25). However, this success has arguably reshaped the dying population. Patients now living with ICDs and advanced structural heart disease constitute a cohort at exceptionally high risk for recurrent, in-hospital monitored arrhythmias (26,27). When these patients experience a terminal electrical event, it frequently occurs during an admission for acute decompensation, post-procedural recovery, or while awaiting advanced therapies such as transplantation or mechanical circulatory support (28). As a result, the very effectiveness of contemporary life-prolonging therapies has inadvertently anchored the most vulnerable patients to hospital-based care at the end of life (29). The modest rise in mortality between 2018 to 2021 may further reflect a growing frail and comorbid cohort that progress to advanced heart disease, where refractory arrhythmias culminating in in-hospital death are more common (30).

We additionally observe a clear divergence in end-of-life courses that tracks closely with patient age and clinical context. Younger adults (25-55 years) and middle-aged adults (45-64 years) were substantially more likely to die in outpatient/emergency department settings and had considerably lower odds of hospice or nursing home death. This pattern aligns with the “first, sudden, and catastrophic” presentation of VA, often as the sentinel manifestation of undiagnosed cardiomyopathy or channelopathy (31,32). These deaths are frequently unheralded, occurring in the community with little opportunity for preemptive palliative care integration. In dramatic contrast, the oldest-old (≥85 years) had significantly higher odds of dying at home or in a hospice/nursing facility. This suggests a different terminal pathway, one where VA may be a final common rhythm in the setting of advanced multi-morbidity and frailty, potentially following a decision to deactivate devices or forego aggressive resuscitation (33,34,35). The near absence of hospice deaths in those under 45 tells us that we rarely involve hospice for young arrhythmia patients. While often appropriate, this highlights that we must not overlook palliative care just because a patient is young, particularly for those with advanced refractory conditions who have exhausted therapeutic options (36). This divergence highlights that age-related clinical profiles and goals of care are central in determining the overall pattern of VA mortality (18).

However, our findings extend beyond clinical heterogeneity to reveal profound and troubling disparities in the setting of VA death, consistent with systemic inequities. Specifically Black, American Indian, and Asian/Pacific Islander decedents had significantly lower odds of home and hospice/nursing facility death compared with White patients. These differences persisted even after adjusting for age, sex, and geographic region, implicating barriers to access, cultural factors, and implicit bias as important determinants. The origins of these disparities are multifactorial and likely parallel those observed in other serious conditions, including longstanding mistrust of the healthcare system, cultural and spiritual meanings attached to death and intensive treatment, and lower frequency of advance care planning discussions (37,38,39). The finding that racial and ethnic minorities were more likely to die in outpatient/ED settings additionally suggests possible delays in seeking care, less availability of bystander resuscitation or disparities within the pre-hospital chain of survival (40,41). Meaningful progress will depend on targeted, culturally informed strategies, including the use of community health workers and value-concordant communication about hospice options (42). We also identified marked sex-based differences, with women exhibiting lower odds of outpatient/ED death and higher odds of hospice/nursing home death, which may reflect differences in VA etiology and symptom profile, as well as social factors such as older age, higher rates of widowhood, and a greater tendency to opt for less aggressive end-of-life care (43,44,45). Geographic gradients were more modest but indicated that rural patients had somewhat higher odds of home death, a pattern that likely reflects limited access to emergency services and hospice infrastructure rather than a pure reflection of patient choice (46,47).

### Implications for Clinical Practice and Policy

Collectively, these findings exert immediate and important influence on clinical practice patterns within electrophysiology and cardiovascular medicine. First, they underscore the pivotal role of proactive, nuanced advance care planning (ACP) for patients with structural heart disease who are at risk for VA. The very limited hospice involvement (**<**6%) indicates a substantial gap in embedding palliative care within electrophysiologic practice. This is particularly critical for the management of implantable devices. Prior work demonstrates that nearly one in three ICD carriers experiences painful shocks during their final days when deactivation is not pursued. Accordingly, proactively framing ICD deactivation as a compassionate pathway to a natural death is crucial for those living with irreversible or terminal disease (48). Electrophysiologists should partner with palliative care teams to manage advanced heart failure symptoms and to facilitate these complex conversations, aiming to ensure that a patient’s terminal arrhythmia is peaceful rather than traumatic.

Second, our analysis offers a strong call to action to confront the disparities we have uncovered. This will demand the adoption of culturally sensitive communication strategies, equipping clinicians with training in shared decision-making, and strengthening community-based outreach. On the policy front, the marked underutilization of hospice indicates the necessity of stronger palliative care integration within cardiology. Possible approaches include implementing automatic palliative care consultation triggers for patients hospitalized with life-threatening arrhythmias and securing robust reimbursement support for advance care planning conversations. Policymakers and health systems should also promote the use of Physician Orders for Life-Sustaining Treatment (POLST) for high-risk patients and establish EMS protocols that honor out-of-hospital DNR orders, thereby reducing non-beneficial transports and enabling death to occur in the patient’s preferred setting.

## Limitations

This study’s key strengths lie in its enormous, nationally representative sample and its 25-year span, which offers an uncommon longitudinal perspective. However, several limitations must be recognized. Our use of death certificate data carries the risk of misclassification of both underlying cause and place of death. For instance, individuals who die at home while enrolled in hospice are recorded simply as “home” deaths, which means that our “hospice facility” designation underestimates total hospice engagement. Furthermore, we lacked granular data on crucial clinical confounders, most notably ICD implantation status, the etiology of the underlying heart disease, and whether device deactivation had occurred prior to death. We also could not capture patient preferences or the quality of the dying experience (e.g., whether an inpatient death was a technology-driven ICU death or a peaceful event). As an ecological study, this analysis reveals population-level relationships yet is not equipped to determine the specific causal pathways that produced these findings.

## Conclusion

In conclusion, this 25-year analysis presents a complex narrative of progress tempered by enduring obstacles in ventricular arrhythmia mortality. The pronounced decline in overall deaths reflects the notable triumphs of modern electrophysiology. Yet, the simultaneous concentration of death within hospitals and the substantial inequities in place of death, underscore a critical frontier for improvement. Advancing compassionate care for patients vulnerable to VA will mean shifting from a singular focus on survival to a broader paradigm that equally prioritizes the quality of death. This will necessitate the electrophysiologists to steer complex conversations about goals of care, work to secure equitable availability of palliative support, and in ensuring that the ICD’s technological sophistication is directed not solely at extending life, but at facilitating an end of life that is aligned with what patient’s value and desire.

## Data Availability

All data produced for this study is publicly available at CDC wonder website.

## Acknowledgements

none

## Author Contributions

**Conceptualization:** M.A.N, A.A,M.O.R; **Data curation:** M.A.N, A.A, M.O.R **Formal Analysis:** M.H, M.J.R; **Methodology:** M.J.R; **Project administration:** M.A.N, M.H, M.J.R **Validation:** M.A.N, A.A, M.O.R; **Visualization:** M.H, M.J.R; **Writing – original draft:** M.A.N, M.J.R, M.H, A.A; **Writing – review & editing**: M.A.N,M.J.R,M.H,A.A,M.O.R.

## Abbreviations

VA: Ventricular Arrhythmia
APC: Annual Percentage Change
AAMR: Age-Adjusted Mortality Rate
CDC WONDER: CDC Wide-ranging Online Data for Epidemiologic Research
SCD: Sudden Cardiac Death
POD: Place Of Death

